# Childcare practices associated with stunting among children aged 6-59 months in agro-pastoral society of Izazi ward of Iringa region

**DOI:** 10.64898/2026.01.15.26344178

**Authors:** Jesca Gaudence Mlay, Maseke Richard Mgabo, Ladigracia Lyakurwa, Alex Fortunatus Magufwa

## Abstract

Child stunting is still a global public health challenge, with a greater impact in developing countries. In Tanzania, an estimate of 30% of children are stunted. This study estimated childcare practices associated with stunting in the agro-pastoral community of Izazi ward in Iringa District. A cross-sectional study was employed to collect both quantitative and qualitative anthropometric data from 377 mother-child pairs of children aged 6-59 months. Three focus group discussions were carried out with mothers, and interviews were conducted with community leaders. Respondents were selected using systematic and snowball sampling methods. Data collection involved a structured questionnaire, where ENA for SMART software was used to generate the index for height-for-age. The findings confirm that stunting is still a public health problem among agro-pastoral communities of Izazi ward, 34.7% of respondents were stunted. A multivariate binary logistic regression was used to assess the childcare practices associated with stunting; factors associated with stunting included: being female (AOR=0.66; 95% CI:0.40-0.99, p=0.048), breastfeeding for 1 year (AOR=19.75; 95% CI:8.99-43.39, p=0.001), breastfeeding for less than two years (AOR=7.08; 95% CI:4.03-12.45, p=0.001), non-exclusive breastfeeding (AOR=2.599; 95% CI:1.23-5.47, p=0.012), and having one disease (AOR=5.36; 95% CI:1.88-15.27, p=0.02). This study highlights the importance of promoting exclusive breastfeeding and treating childhood illnesses to improve health outcomes among children.

## INTRODUCTION

Stunting is still a public health concern that affects children below the age of five. It is defined as a low height for age Z-score of the reference population reflecting undernutrition. According to the World Health Statistics report of 2022, 148.1 million children are approximately to be stunted. Several factors such as food and nutrition crises, conflicts, climate change, and the COVID-19 pandemic were attributed to stunting [1].

The African continent has the highest stunting rates globally, as it is estimated to be 30%, which is significantly higher than the global estimate of 22.3% [2]. This rate is attributed to food insecurity, low living standards, poor health care services, and climatic disasters such as droughts and floods [3,4].

In agro-pastoral communities, where economic activities include crop cultivation and livestock keeping, livestock are vulnerable to stunting due to an unstable climate and economic hardship [5].

The prevalence of stunting varies by region, with the highest observed in African regions, where 56.2 million children under five are stunted, followed by 49.8 million children under five in South East Asia and 22.9 million children in Eastern Mediterranean regions [1].

In Tanzania, the prevalence of stunting is estimated to be 30% [6]. The prevalence also varies by region, with the highest prevalence registered in Iringa (57%), Njombe (50%), Rukwa (50%), Geita (39%), and Simiyu (33%) regions. While the lowest prevalence is registered in Mjini Magharibi (13%), Kusini Pemba (17%), Kusini Unguja(17%), Dar es Salaam 18%, Kilimanjaro 20%, and Pwani 20% regions.

According to a 2023 FAO report, poor nutrition and maternal health, inadequate feeding practices for both youth and young children, and diseases were the causes of stunting [1]. Furthermore, rural areas of semi-pastoral communities are mostly affected due to poor health services and inadequate food sources. The report further indicates that factors contributing to this include the mother’s education, inadequate dietary diversity, high rates of infectious diseases, and economic hardships [5].

The Tanzanian government has taken several initiatives for decades to combat malnutrition through various strategies such as Work for Food program, Food subsidies, school feeding, Most Vulnerable Children’s program [7]. The recent effort has been the formation of the National Multisectoral Nutrition Action Plan (NMNAP) 2016-2021. This common framework tracks collective progress to end malnutrition in Tanzania [8].

Iringa has been performing relatively poorly ever since 1992 [9]. Some organisations like GAIN and Global Volunteers have made different efforts to combat stunting in the region through initiatives such as food fortification, establishing health clinics, and implementing the Reaching Children’s Potential (RCP) program [10–12]. However, the prevalence of stunting remains unacceptably high 57% [6]. Despite the efforts made by the government and other partners to combat stunting in the Iringa region; stunting is still a health challenge. This study, therefore, explored childcare practices associated with stunting among children aged 6-59 months in the study area.

## Materials and Methods

### Research design and study area

A community-based cross-sectional study was conducted in Izazi ward of Iringa District, Southern Tanzania, from 17 February to 17 April 2025. Both qualitative and quantitative methods were used in data collection. While Izazi ward was selected among the 20 wards of Iringa district due to its agro-pastoral features. The three villages of Izazi ward (Izazi, Makuka, and Mnadani) were the targeted area for the study. It lies between latitudes 7° 12’ 0” South, 35° 44’ 0” East. It is accessible via Highway A104, approximately 27 miles north of the main town of Iringa [13]. According to the 2022 census, Izazi ward had a total population of 8764. Izazi is predominantly an agro-pastoral community where the economy is mainly depend on livestock farming and crop cultivation. Herding and animal husbandry play central roles in the livelihoods of its residents, agricultural activities playing a minor role in the area. Crops cultivated are mainly sunflower and, in small amounts of maize and millet, livestock keeping mainly cows, goats, and sheep [14]. There are low standards of living and poor sanitation practices.

### Targeted population

Mother-child pairs of children aged 6-59 months were the targeted population.

### Study Population

This study comprises children aged 6-59 months and their mothers in the sampled household, in a household with more than one child aged 6-59 months; only one child was selected to represent others in a household. *Inclusion criteria:* mother-child pairs of children aged 6-59 months who were not critically sick during the survey were involved in the study. *Exclusion criteria:* mother-child pairs of children aged 6-59 months who are critically sick during the survey were not included in the study, as this could bring difficulties in stunting measurement.

### Sample size and sampling

#### Sample size

The sample size was calculated using the assumption 95% confidence interval, the proportion of stunting in the Iringa region of 57%, and 5% margin of error. 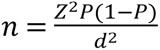[15].

where n = sample size, Z = Z statistic for a level of confidence (for the level of confidence of 95%, which is conventional, Z value is 1.96), P = expected prevalence or proportion (prevalence of stunting in Iringa is 57%, P = 0.57), and d = allowable error (d = 0.05).

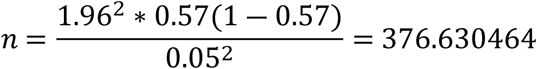

The total sample size was 377.

### Sampling procedures

A proportionate sampling technique was employed to select participants from target villages within the ward. To obtain mothers with children aged 6-59 months in a village, a systematic sampling method was applied using the health records of a targeted village. For the mothers who do not attend clinics, a snowball method was applied to get them. In a household with more than one child aged 6-59 months, only one child was selected to represent others in a household. In addition, a purposive sampling technique was used to select key informants, such as health workers and the village health committee, the District Nutritionist, Village Health Community Officer.

### Data collection methods and tools

A questionnaire developed by researchers was used to collect data from mother-pairs of children aged 6-59 months on socio-demographic characteristics of the household, child feeding practices and healthcare practices associated with stunting in their area of residence.

#### Assessment of stunting

In assessing the nutritional status of children, anthropometric measurements of height, weight, and age were used. The height of infants aged between 6 months and 23 months was measured in a recumbent position to the nearest 0.1 cm using a board with an upright wooden base and a movable headpiece. Children between 24 and 59 months of age were measured in a standing position to 0.1 cm to the nearest. Parents were asked the age of their children, and was confirmed from birth certificates and clinic cards, for those who did not remember their child’s age probing technique was used to get the child’s age. A weighing scale was used to measure the weight of all children to the nearest 0.1 g.

#### Qualitative data collection

Three Focus group discussions, each comprising a minimum of 6 and a maximum of 8 self-selection participants, were conducted during clinic-hosted health education programs on nutrition education and feeding practices. The researcher facilitated the discussions by guiding participants through the topics sequentially. The topics covered included child feeding practices and healthcare practices and participants were encouraged to express their views freely.

### Ethical consideration

Ethical clearance was obtained from the Institute of Rural Development Planning. A formal letter was written to different administrative bodies to obtain permission and resources to conduct the research in the setting. The agro-pastoral households with children aged 6-59 months were informed the purpose, benefit, and assured confidentiality. Thereafter, verbal consent was obtained from them. Participation in the study was voluntary, and participants were allowed to withdraw at any time from the study and were allowed to ask study-related questions.

### Quality control

The questionnaire was prepared in English, translated into Swahili, and returned to English to maintain consistency. A pre-test of the questionnaire was done before the actual data collection to estimate the time needed, and some modifications were made based on the pre-test findings. The anthropometric measurement tools were tested before the actual measurement to ensure the consistency of the tools. In addition, an introduction was given to respondents on the aim of the study before data collection.

### Data Entry and Analysis

#### Quantitative data

The collected data of Sex, weight, height, and age were entered into ENA for SMART software using the WHO standards 2006 with participant identification numbers to translate nutritional data into Z scores for the HAZ indices. Finally, the data, including the HAZ, were exported to STATA software version 17.0 for analysis. The binary logistic analysis was performed to determine the childcare practices that were associated with stunting. The variables that were associated at p-value<0.2 were selected for multivariable binary logistic regression. Finally, the variables that showed a significant association were selected based on the adjusted odds ratios (AOR) with the 95% confidence interval and p-value<0.05.

#### Qualitative data

The researchers used a thematic approach in analyzing data, whereby raw data in the form of recordings and notes were arranged for easy access and review, then were familiarized by the researchers to understand the context, and then coded to present key concepts and ideas of the data. Finally, data were presented in the form of quotations to represent ideas from the corresponding respondents.

## Results and Discussion

### Social demographic factors associated with stunting

As indicated in Table 1, female children reduced the stunting rate by 70%. Children aged 12-23, 24-35 and 36-47 months had Odds Ratio 1.60, 1.52 and 1.33 times respectively more likely to be stunted compared to those aged 6-11months and 48-59 months. During FGDs income generating activities like farming, business and milking were revealed as reason for reduced child caring among child aged above 1 year leaving the responsibility to their older sibling, which affects timely feeding nutrition adequacy and health monitoring of children, as revealed by 1 mother from the Maasai community. Their narrations revealed mothers spend less time with their children compared to their activities.

> *“those of us who have stopped breastfeeding we usually wake up early to go sell milk in the village, leaving our children with their older siblings, due to the distance between here (homes) and the village, we usually return in the afternoon to cook lunch for them”.*

**Table 1:** Logistic regression of the association between socio-demographic factors and stunting.

| **Variable** | **Stunted**<br>**HAZ<-2SD**<br>**n(%)** | **Not stunted**<br>**HAZ≥ -2SD**<br>**n(%)** | **Crude Odds Ratio**<br>**(95% CI)** | **P-values** |
| --- | --- | --- | --- | --- |
| **Child's age in months** |  |  |  |  |
| 6-11 | 15(4.0) | 34(9.0) | 1.00(reference) |  |
| 12-23 | 32(8.5) | 50(13.3) | 1.605(0.758-3.398) | 0.216 |
| 24-35 | 36(9.5) | 56(14.9) | 1.525(0.729-3.185) | 0.262 |
| 36-47 | 29(7.7) | 52(13.8) | 1.333(0.625-2.841) | 0.456 |
| 48-59 | 19(5.0) | 54(14.3) | 0.797(0.357-1.777) | 0.580 |
| **Child's gender** |  |  |  |  |
| Male | 77(20.40) | 120(31.80) | 1.00(reference) |  |
| Female | 54(14.30) | 126(33.40) | 0.707(0.462-1.080) | 0.109 |
| **Birth weight** |  |  |  |  |
| less than 2.5 | 4(1.1) | 5(1.3) | 1.00(reference) |  |
| 2.5-3.5 | 120(31.8) | 227(60.2) | 0.695(0.183-2.635) | 0.593 |
| greater than 3.5 | 5(1.3) | 13(3.4) | 0.481(0.090-2.556) | 0.390 |
| Not known(born home) | 2(0.5) | 1(0.3) | 2.5(0.162-38.599) | 0.512 |
| **Child's birth interval** |  |  |  |  |
| 1 year | 91(24.2) | 160(42.6) | 1.00(reference) |  |
| 2 years | 38(10.1) | 75(19.9) | 0.837(0.525-1.334) | 0.455 |
| more than 2 years | 2(0.5) | 10(2.7) | 0.331(0.071-1.541) | 0.159 |
| **Household income/day** |  |  |  |  |
| 1000-5000Tsh | 72(19.1) | 138(36.6) | 1.00(reference) |  |
| 6000-10000Tsh | 54(14.3) | 100(26.5) | 1.072(0.694-1.655) | 0.752 |
| More than 10000Tsh | 5(1.3) | 8(2.1) | 1.608(0.521-4.963) | 0.408 |
| **Household size** |  |  |  |  |
| 1-3 members | 16(4.20) | 26(6.90) | 1.00(reference) |  |
| 4-6 members | 84(22.30) | 167(44.30) | 0.674(0.321-1.412) | 0.296 |
| >6 members | 31(8.20) | 53(14.10) | 0.701(0.293-1.680) | 0.427 |

Another mother from Sukuma community added

> *“We Sukuma, we live in polygamous marriage, you find one husband has 2 to 4 wives and our main occupation is farming, so during farming season one wife with the youngest child is assigned to stay home and take care of all the young children while we are in the field until we return in the evening”.*

Over three-quarter of children (92%) were born with weight between 2.5-3.5 kg, about two-third of children (66.8%) had birth interval of 1 year. Over half of mothers (56%) had no formal education. During FGDs it was revealed that, female children of school age were responsible for taking care of their young siblings when their mothers were not around or gone for livestock rearing, causing more female children to grow up without schooling. One mother said “*Back then, my farther had many cattle, so my siblings and I were responsible for finding the cattle’s fodder. However, nowadays male children are in charge of raising livestock, while female children look after their young children*”.

Two-third of children (66.6%) are from household with 4-6 members and more than half of households (55.7%) gain 1000-5000 Tsh per day.

### Child feeding practices associated with stunting

As indicated in Table 2, Over half of children (53.3%) were initiated breastfeeding within the first hour after delivery, over two-third of children (71.6%) were not exclusively breastfed for the first six months, over half of children breastfed for more than 2 years. During FGDs most mothers revealed the difficulties of practicing exclusive breastfeeding as one mother revealed “Haaaa*! How can do exclusive breastfeeding for 6 months? I used to prepare maize porridge for my children when they are two months old. It is easier because a baby get fed-up and gives you time to do other activities*” (2 mother). A mother of three children added,

> *“To my side, breast milk is a problem. I might stay up to two days after birth with no milk, so my children are initiated with sugar or glucose since day one, and I see it as normal.”(Lamented mother 3)*

**Table 2:**
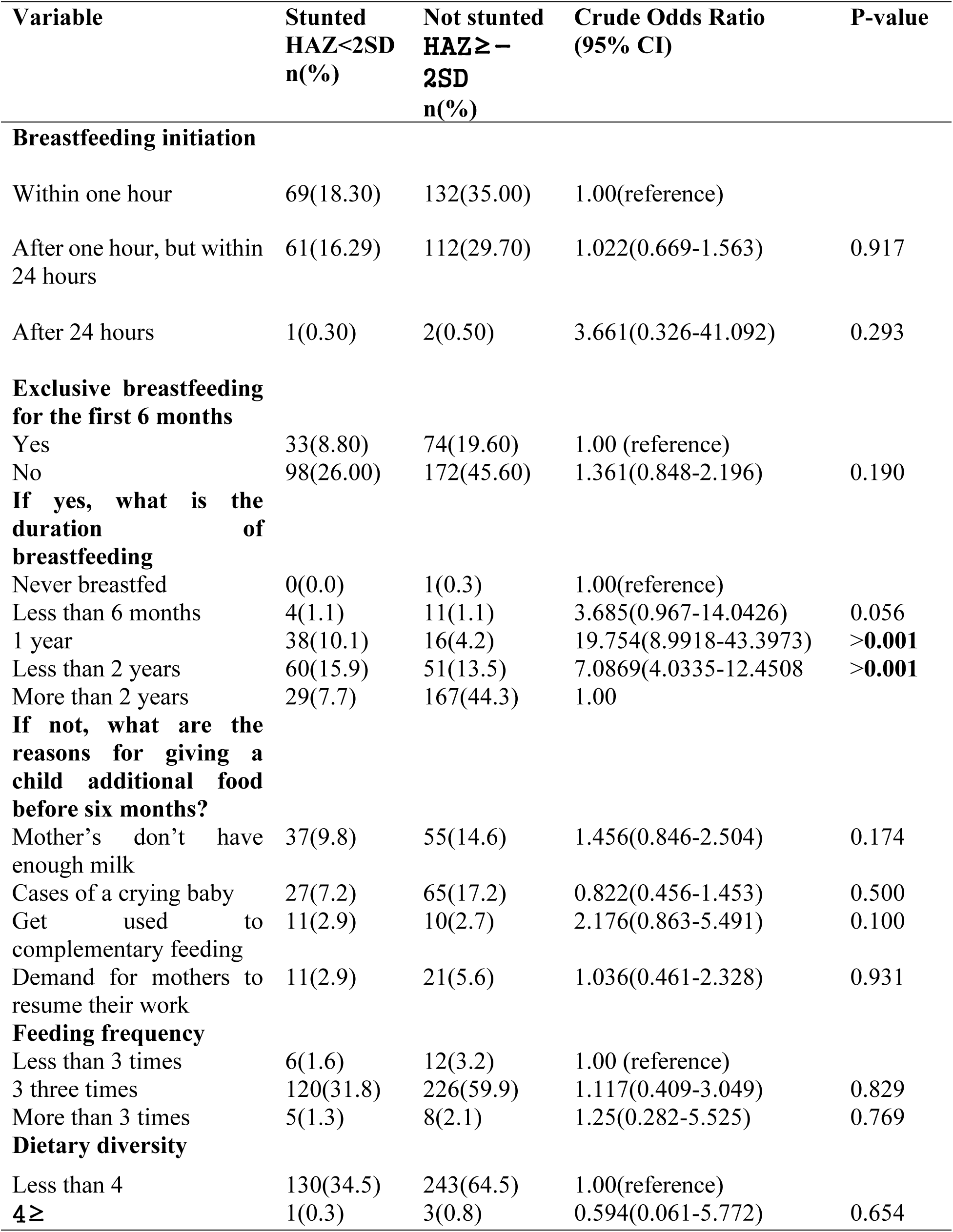
Logistic regression analysis of child feeding practices associated with stunting.

More than three-quarter of children (91%) were fed three times per day and majority of children (98.9%) had less than 4 dietary diversities in the prior to survey. In the focus group discussion, some respondents reported that food is prepared once per day and eaten from morning to evening,

> *“ Our staple food is maize (carbohydrate) this is main [maize] with variety of sauces like beans, peas, fish, vegetables, sometimes meat, in most cases sauces is unproportioned to a recommended portion as nurses teach us” ( Reported a mother 4)*

> *“Members are heavily reliant on millet, maize, sesame crops, and livestock keeping, with limited access to fruits and legumes, hence influencing low nutritional diversity. In terms of vegetables, we eat vegetables throughout the year, during dry season we eat stocked dried vegetables” … Some mother even feed their babies with cold leftovers (A key Informant, community development officer lamented)*

### Healthcare practices associated with stunting

According to Table 3, over two-thirds of children (69.2%) were fully vaccinated and over quarter were partially vaccinated. During Key Informant Interview, the findings revealed a wide coverage of vaccine in the area, hence partial status was due to timing of vaccine dosses rather than missed or refused vaccination “*In our setting the immunization coverage has achieved coverage for the targeted age group, especially children aged 12-23 months, through regular outreach and fixed site sessions*” Ward Executive Officer.

**Table 3:**
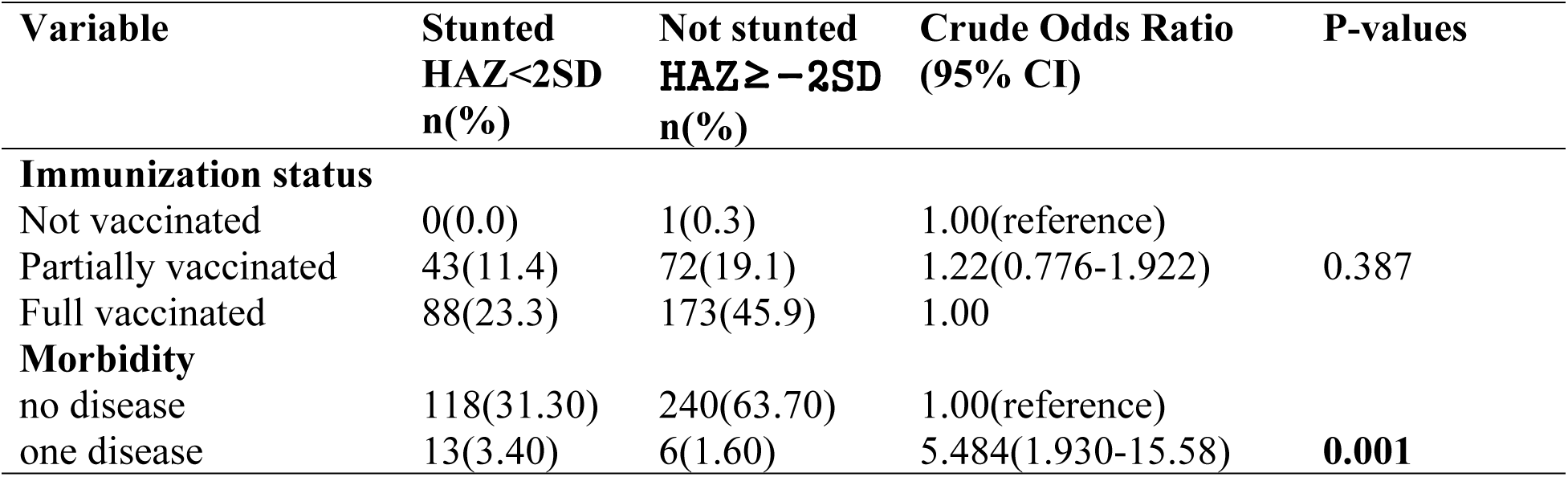
Logistic regression analysis of Health care practices associated with stunting.

Over three-quarter of children (95%) reported no presence of disease in two weeks prior to survey.

### 3.4 Prevalence of stunting

The prevalence of stunting was found to be 34.7% (26.8-43.7 95%CI). In addition, the prevalence of severe and moderate stunting was 13.0% and 21.8% respectively. The prevalence of stunting among boys and girls was 39.1% and 30.0% respectively, Table 4.

**Table 4:** Prevalence of stunting based on height-for-age Z-scores and by sex.

|  | **All**<br>n = 377 | **Boys**<br>n = 197 | **Girls**<br>n = 180 |
| --- | --- | --- | --- |
| **Overall stunting(<-2 z-score)** | (131) 34.7 % | (77) 39.1 % | (54) 30.0 % |
| **Prevalence of moderate stunting (<-2 z-score and >=-3 z-score)** | (82) 21.8 % | (48) 24.4 % | (34) 18.9 % |
| **Prevalence of severe stunting (<-3 z-score)** | (49) 13.0 % | (29) 14.7 % | (20) 11.1 % |

The highest prevalence of stunting was 40.8% among children aged 24-35months, 37.4% among children aged 12-23 months, 35 9% among the age group of 36-47 months, 28.6% among children aged 6-11 months, and the lowest was 27.1% among children aged 48-59 months. (Fig 1)

**Figure 1:**
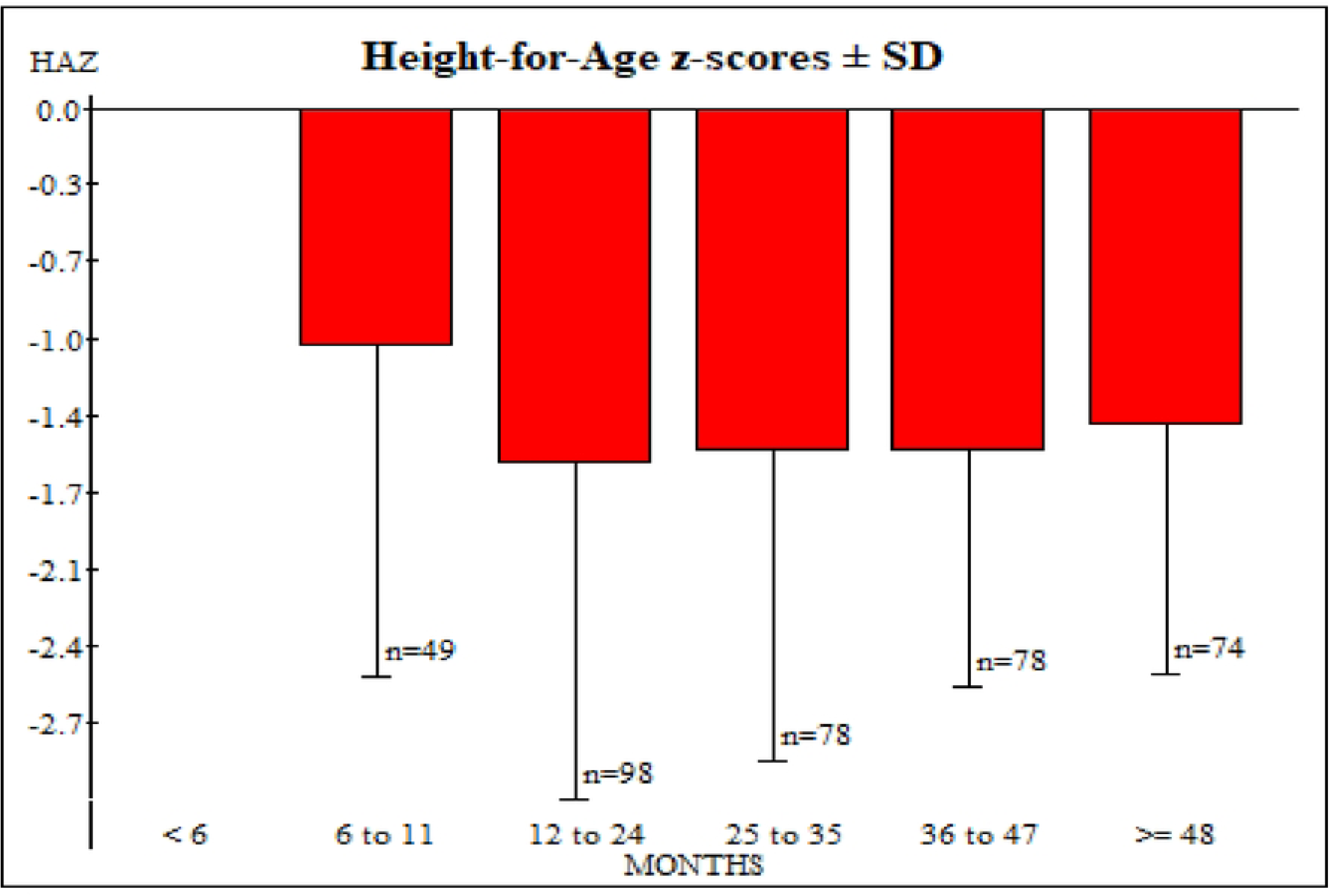
Height-for-age Z-scores based on age Childcare practices associated with stunting

### Childcare practices associated with stunting

According to results in Table 5, Stunting was associated with exclusive breastfeeding, child’s gender, morbidity and duration of breastfeeding. Children who were non-exclusively breastfed for the first six months were 2.59 times more likely to be stunted compared to those who were exclusive breastfeeding for the first six months (AOR=2.599; 95% CI:1.23-5.47, p=0.012). Duration of breastfeeding was associated with increased stunting rate especially to children who breastfed for 1 year (AOR=19.75; 95% CI:8.99-43.39, p=0.001) and less than 2 years (AOR=7.08; 95% CI:4.03-12.45, p=0.001) respectively.

**Table 5:** Multivariable analyses of variables associated with stunting in children aged 6-59 months in agro-pastoral community of Izazi ward.

| **Variable** | **Stunted**<br>**HAZ<**<br>**2SD**<br>**n(%)** | **Not**<br>**stunted**<br>**HAZ≥**<br>**-2SD**<br>**n(%)** | **Crude Odds**<br>**Ratio**<br>**(95% CI)** | **P-**<br>**value**<br>**s** | **Adjusted**<br>**Odds Ratio**<br>**(95% CI)** | **P-**<br>**value**<br>**s** |
| --- | --- | --- | --- | --- | --- | --- |
| **Exclusive**<br>**breastfeeding** |  |  |  |  |  |  |
| Yes | 33(8.80<br>) | 74(19.6<br>0) | 1.00<br>(reference) |  |  |  |
| No | 98(26.0<br>0) | 172(45.<br>60) | 1.361(0.848-2.196) | 0.190 | 2.599 (1.233-5.479) | **0.012** |
| **Child's**<br>**gender** |  |  |  |  |  |  |
| Male | 77(20.4<br>0) | 120(31.<br>80) | 1.00(referenc<br>e) |  |  |  |
| Female | 54(14.3<br>0) | 126(33.<br>40) | 0.707(0.462-1.080) | 0.109 | 0.660(0.404-0.996) | **0.048** |
| **Morbidity** |  |  |  |  |  |  |
| no disease | 118(31.<br>30) | 240(63.<br>70) | 1.00(referenc<br>e) |  |  |  |
| one disease | 13(3.40<br>) | 6(1.60) | 5.484(1.930-15.58) | **0.001** | 5.369(1.886-15.279) | **0.002** |
| **Duration of**<br>**breastfeeding** |  |  |  |  |  |  |
| Never<br>breastfed | 0(0.0) | 1(0.3) | 1.00(referenc<br>e) |  |  |  |
| Less than 6<br>months | 4(1.1) | 11(1.1) | 1.796(0.538-5.987) | 0.340 | 3.685(0.967-14.0426) | 0.056 |
| 1 year | 38(10.1<br>) | 16(4.2) | 11.731(5.862<br>-23.474) | **0.001** | 19.754(8.9918<br>-43.3973) | **0.001** |
| Less than 2<br>years | 60(15.9<br>) | 51(13.5<br>) | 5.811(3.425-9.857) | **0.001** | 7.0869(4.0335<br>-12.4508) | **0.001** |
| More than 2<br>years | 29(7.7) | 167(44.<br>3) |  | 1.00 | 1.00 |  |
| **Child's birth**<br>**interval** |  |  |  |  |  |  |
| 1 year | 91(24.2<br>) | 160(42.<br>6) | 1.00(referenc<br>e) |  |  |  |
| 2 years | 38(10.1<br>) | 75(19.9<br>) | 0.837(0.525-1.334) | 0.455 | 0.887(0.536-1.468) | 0.64 |
| more than 2<br>years | 2(0.5) | 10(2.7) | 0.331(0.071-1.541) | 0.159 | 0.259(0.455-1.481) | 0.129 |

Children who were sick with 1 disease 2 weeks prior to survey had 5.36 times of being stunted compared to children with no disease (AOR=5.36; 95% CI:1.88-15.27, p=0.02). Female children were 0.66 times less likely to be stunted compared to male children (AOR=0.66; 95% CI:0.40-0.99, p=0.048).

## Discussion

The prevalence of stunting among children aged 6-59 months in the current study was 34.7%, the prevalence of severe and mild stunting was 13.0% and 21.8%, respectively. The level of stunting in this study is specified to be very high in the study area, according to WHO classifications [16]. According to the Tanzania Demographic and Health Survey (TDHS) report of 2022, the estimated stunting rate is about 30% but prevalence in this study is slightly higher than the national average, highlighting the ongoing nutritional issues in rural and agro-pastoral settings. Comparing this prevalence with other regions in Tanzania and other developing countries, the prevalence was lower than the study conducted in Morogoro, Tanzania, which found a prevalence of 54.3% (Mohamed and Nyaruhucha 2022), and Northern Ethiopia, 39.5% [17]. Nevertheless, the prevalence in the current study area was higher than the studies conducted in Zambia, 21% [18], Mexico, 12.3% [19], Longido Arusha, Tanzania, 31.6% [5], and in Uganda, 22.37% [20].

Besides, stunting before the age of two is forecasted to have poor cognitive and educational outcomes in childhood and adolescence, which may later result in non-communicable diseases such as diabetes and heart disease in adulthood. This later may affect some of labour productivity, income-earned potential, and social skills [2]. Thus, eliminating stunting is vital for family and national prosperity.

The current study indicates that children who were not exclusively breastfed for the first 6 months after birth were more likely to be stunted compared to those who were exclusively breastfed for the first 6 months. This could be because most of mothers do not have enough knowledge and awareness on the importance of exclusive breastfeeding for the first six months. In addition, activities such as income-generating activities, like farming and livestock products businesses, kept some mothers busier than taking care of their children. This is in line with other studies conducted in Ethiopia [17,21], Uganda [20].

The current findings revealed that female children were less stunted compared to male children. This study is in line with other literature that revealed male children were more stunted compared to female children due to sex preferences of the family [21–23]. The reason behind this could be that boys are hungrier compared to girls as they are introduced to early weaning [24]. Although some literature showed no significant difference in stunting between boys and girls [25].

Children with at least one disease such as diarrhoea, respiratory diseases, and malaria, during two weeks before the day of survey were more likely to be stunted. The probable explanation is that repeated diseases weaken the body immune system as the energy that were to be used in growth is used in different metabolic activities to fight against diseases hence impair normal body growth. This finding is in line with several studies [17,21].

Shorter duration of breastfeeding was associated with increased stunting in the current study. This finding is in line with the study conducted in Tanzania [26], Mexico [19], and Indonesia [27] which reported insufficient and inappropriate breastfeeding are correlated with higher stunting rates among children under five.

### Limitation of the study

Recall bias among mothers in answering questions relating to their own age and their children, self-reporting bias was encountered, especially in questions related to income and feeding frequency of a child. In addition, we did not collect data on food security that could increase the prevalence of stunting.

## Conclusion and Recommendations

In conclusion, in the agro-pastoral community, the prevalence of stunting was critically high, suggesting stunting is still a public health concern. The study abled to determine childcare practices associated with stunting, including morbidity, child’s gender, duration of breastfeeding, and exclusive breastfeeding. The government and other partners should promote healthcare and treatment of childhood diseases to reduce stunting, as well as emphasize home gardening to increase access to diverse foods, including fruits and vegetables. At the community level, community development officers can interne through awareness creation programs on cultural shift in social norms to redistribute household unpaid care work to relief women from work lord. Further studies should focus on hygiene and sanitation practices that are associated with stunting in the area.

## Data Availability

all data are publicly available at https://drive.google.com/drive/folders/1Xbp-dbWbz9cV06B25WodrXMqLZL6cr-q?usp=sharing

## Data availability statement

The data is available upon request from the authors.

## Acknowledgement

The authors express their appreciation to the Iringa district Council, Institute of Rural Development Planning, data collectors and all mother-child pair of children aged 6-59 months who participated in the study.

## Author Contributions

**Conceptualization:** Jesca G Mlay, Maseke R Mgabo

**Formal analysis:** Jesca G Mlay

**Investigation:** Jesca G Mlay,

**Supervision:** Maseke R Mgabo, Ladigracia Lyakurwa

**Visualization:**Jesca G Mlay, Alex F Magufwa

**Writing-original draft:** Jesca G Mlay, Maseke R Mgabo, Ladigracia Lyakurwa

**Writing-review& editing:** Jesca G Mlay, Maseke R Mgabo, Ladigracia Lyakurwa, Alex F Magufwa

